# Prediction of stroke-associated pneumonia risk in stroke patients based on interpretable machine learning

**DOI:** 10.1101/2024.10.27.24316222

**Authors:** Chunbiao Li, Ting Wang, Juan Yuan, Linli Yuan, Min You

**Author notes:** **Correspondence:** Ting Wang, MD. These authors contributed equally to this work (Chunbiao Li, Juan Yuan).

## Abstract

**Background:** Stroke-associated pneumonia (SAP) is a frequent complication of stroke, characterized by its high incidence rate, and it can have a severe impact on the prognosis of patients. The limitations of current clinical treatment measures underscore the critical need to identify high-risk factors promptly to decrease the incidence of SAP.

**Objective:** To analyze the risk factors of SAP in stroke patients, construct a predictive model of SAP based on the SHAP interpretable machine learning method, and explain the important variables.

**Methods:** A total of 763 stroke patients admitted to the Second Affiliated Hospital of Anhui University of Traditional Chinese Medicine from July 1, 2023, to May 31, 2024, were selected and randomly divided into the model training set (n=457) and model validation set (n=306) according to the ratio of 6:4. Firstly, the included data were sorted out, and then Lasso regression was used to screen the included characteristic variables. Based on the tidymodels framework, Using decision tree (DT), logistic regression, extreme gradient boosting (XGBoost), support vector machine (SVM), The classification model was constructed by five machine learning methods, including SVM and LightGBM. The grid search and 5-fold cross validation were used to optimize the hyperparameter optimization strategy and the performance index of the model. The predictive performance of the model was evaluated by the area under the receiver operating curve (AUC), calibration curve, and decision curve analysis (DCA), and we used Shapley additive explanation (SHAP) to account for the model predictions and provide interpretable insights.

**Results:** The incidence of SAP in this study was 31.72% (242/763). Six variables were selected by Lasso regression, including nasogastric tube use, age, ADL score, Alb, Hs-CRP, and Hb. The model with the best performance in the validation set was the XGBoost model, with an AUC of 0.926, an accuracy of 0.914, and an F1 score of 0.889. Its calibration curve and DCA showed good performance. SHAP algorithm showed that ADL score ranked first in importance.

**Conclusion:** The model constructed using XGBoost has good prediction performance and clinical applicability, which is expected to support clinical decision-making and improve the prognosis of patients.

## 1. Introduction

Stroke is a severe cardiovascular disease that significantly impacts the quality of life and survival rate of patients. According to the Global Burden of Disease Study, stroke caused approximately 6.55 million deaths in 2019, ranking as the second leading cause of death worldwide, second only to cardiovascular diseases[1]. In the aftermath of a stroke, patients frequently contend with numerous complications, with stroke-associated pneumonia (SAP) being among the most prevalent[2]. Surveys have indicated that the incidence of SAP ranges from 7% to 38%[3–6], SAP not only prolongs hospitalization and increases economic burdens but can also severely affect patient mortality[4–5]. Currently, the primary treatment for SAP in clinical practice is anti-infective therapy [7]; however, studies have shown that prophylactic antibiotics do not effectively reduce the risk of SAP occurrence[8]. Therefore, it is crucial for clinicians to promptly identify high-risk individuals for SAP and implement appropriate preventive measures, which are essential for improving the prognosis of stroke patients.

The establishment of risk prediction models can assist clinicians in identifying high-risk populations for diseases, allowing for early intervention measures to reduce disease incidence[9]. In recent years, scholars both domestically and internationally have developed multiple models to predict the risk of SAP occurrence, presented in the form of scoring systems and scorecards, such as the Kwon Score[10], A2DS2 Score[11], and AIS-APS Score[12]. These models provide relatively reliable assessment tools for the prevention and treatment of SAP. However, even well-validated models may experience performance degradation over time due to changes in disease risk factors, treatment measures, and treatment contexts. Therefore, models need continuous dynamic updates [13]. In addition, few studies are using interpretable machine learning to construct SAP risk prediction models. Based on this, this study considered combining new predictors with known predictors to construct a model using the machine learning method and using the SHAP algorithm to explain the model, to improve the accuracy and interpretability of SAP risk prediction.

## 2. Materials and Methods

### 2.1 Study Design and Subjects

This study is a single-center, retrospective cohort study. We selected stroke patients who were treated at The Second Affiliated Hospital of Anhui University of Chinese Medicine from July 1, 2023, to May 31, 2024, as the study subjects. Inclusion criteria were: (1) patients diagnosed with stroke; (2) age ≥18 years; (3) no mechanical ventilation within 7 days post-stroke. Exclusion criteria were: (1) discharge, transfer, or death within 24 hours of admission; (2) pre-existing pulmonary infection prior to admission; (3) abandonment of treatment or voluntary discharge. (4) the missing rate of collected data exceeded 30%.This study was approved by the Ethics Committee of the Second Affiliated Hospital of Anhui University of Traditional Chinese Medicine (2023-SXXM43). In addition, informed consent was omitted for all participants as the study was retrospective in nature, and the research process was in accordance with the Declaration of Helsinki.

### 2.2 Identification of Candidate Predictive Factors

Candidate predictive factors for this study were identified through literature review and expert consultation, totaling 27 factors, including: (1) General demographic data: gender, age; (2) Disease-related factors: activity of daily living (ADL) scale[14] score at admission, type of stroke, location of stroke, dysphagia, impaired consciousness, hypertension, diabetes mellitus; (3) History of comorbidities/Personal history: history of stroke, history of underlying pulmonary disease, smoking, drinking; (4) Disease treatment factors: nasogastric tube, acid suppressants, urinary catheters; (5) Laboratory test indicators: albumin (Alb), triglyceride (TG), hypersensitive C-reactive protein (Hs-CRP), white blood cell count (WBC), neutrophil-to-lymphocyte ratio (NLR), monocyte-to-lymphocyte ratio (MLR), hemoglobin (Hb).

### 2.3 Definition and Diagnosis of SAP

According to the consensus published by the SAP Consensus Group composed of multidisciplinary experts in the UK[2], the outcome indicator SAP is defined as pneumonia that newly occurs within 7 days in non-mechanically ventilated stroke patients. The diagnosis of SAP follows the modified standards of the Centers for Disease Control and Prevention (CDC) [15].

### 2.4 Sample Size Estimation

This study utilized the sample size estimation method specifically designed for developing risk prediction models, as proposed by Riley et al. in 2020[16]. This method takes into account the effects of multiple categories, interactions, and non-linear relationships, minimizing the risk of model overfitting while precisely estimating key parameters to determine the appropriate sample size required to construct the predictive model. The specific calculations were performed using the “pmsampsize” package in R software. Based on the literature, the average C-statistic of existing prediction models is approximately 0.827[17], and the incidence of SAP is around 7%-38%[3–6]. This study anticipated the inclusion of 27 predictive factors, with the calculated required sample size ranging from 701 to 1272 cases, and an expected number of outcome events ranging from 179 to 253. The detailed calculation process and results are shown in the supplementary material.

### 2.5 Data Collection and Preprocessing

Data sources were determined by reviewing electronic medical records, including admission records, discharge summaries, nursing records, and laboratory results, with laboratory parameters primarily collected from the first admission data. During data collection, we ensured blinding between the predictive factors and outcome indicators to avoid information bias. To address missing data issues, the random forest method was used for data imputation to maintain data integrity and the accuracy of the predictive model (imputation of missing values was performed using the “mice” package in R software, with five imputations). To prevent information loss, all continuous variables were inputted using their original values; for categorical variables, categories with low proportions were considered for merging. After data processing, the dataset was divided into training and validation sets in a 6:4 ratio. The training set data were used to fit the predictive model, while the validation set data were used to evaluate the model.

### 2.6 Model construction and evaluation

Taking the occurrence of SAP in stroke patients as the dependent variable and alternative predictors as the independent variable, least absolute shrinkage, and selection operator (Lasso) regression was used to screen the variables, and then a model was constructed based on the tidymodels framework. We selected the following five machine learning algorithms to build the model: decision tree (DT), logistic regression (LR), extreme gradient boosting (XGBoost), support vector machine (SVM), and light gradient boosting machine (LightGBM). A 5-fold cross-test was performed on the training set as a resampling method, and the hyperparameters were optimized by grid search. We used the area under the receiver operating characteristic (ROC) curve (AUC) to evaluate the discrimination of the model and drew calibration curves to evaluate the fit of the model. Decision curve analysis (DCA) was used to evaluate the clinical practicability of the model. Finally, the accuracy, sensitivity, specificity, and other indicators of the model were calculated to evaluate the performance of the model.

### 2.7 Interpretability of the model

Shapley additive explanation (SHAP) is a popular method of model explanation, which is based on the Shapley value in game theory and is used to explain the predictions of any model. The core idea of SHAP is to decompose the contribution of each feature to the prediction, so that the model prediction can be expressed as a weighted sum of feature contributions, thereby improving the transparency and trust of the model and supporting more reasonable decision making. By applying the SHAP algorithm to the optimal model, we can obtain the importance ranking of features and have an intuitive understanding of the contribution of these features to the prediction model.

### 2.8 Statistical Methods

Statistical analysis was performed using R 4.3.0 software.Normally distributed continuous data were expressed as mean ± standard deviation (*Mean* ± *SD*), and between-group comparisons were performed using the independent sample t-test. Non-normally distributed continuous data were expressed as median and interquartile range [*M* (*Q*1, *Q*3)], and between-group comparisons were conducted using the Mann-Whitney U test. Categorical data were expressed as counts and percentages [*n* (%)], and comparisons of unordered categorical data between groups were performed using the Pearson x^2^ test or Fisher’s exact test. All statistical tests were two-sided, with *P*<0.05 considered statistically significant.

## 3. Results

### 3.1 Missing Data

There were 7 variables with missing values in this study, including Alb, TG, LDL, HDL, Stroke location, Hs-CRP, and D-Dimer. The missing values were all imputed by the random forest method (Fig 1).

**Fig 1.**
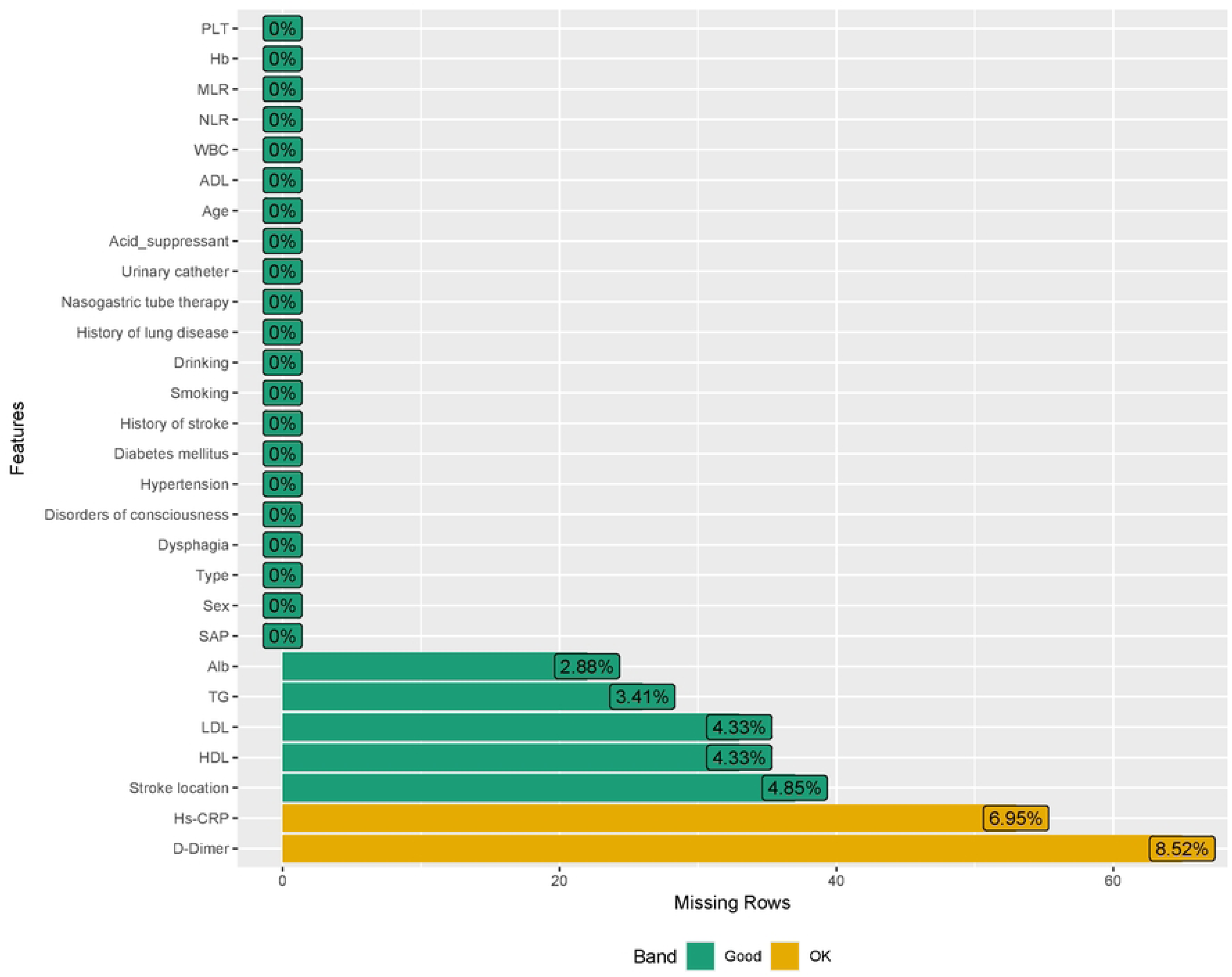
Missing data.

### 3.2 Patient Characteristics

Based on the occurrence of SAP, the 763 patients were divided into the non-SAP group (*n*=521) and the SAP group (*n*=242). Among the 763 included patients, there were 504 males (66.06%) and 259 females (33.94%), with an average age of 67.35±13.00 years. The incidence of SAP was 31.72%. There were statistically significant differences in age, ADL score, type of stroke, dysphagia, disturbance of consciousness, diabetes, history of lung disease, nasogastric tube therapy, urinary catheter, Alb, Hs-CRP, WBC, NLR, MLR, Hb, D-Dimer and HDL between the two groups (*P* < 0.05). The detailed results are presented in Table 1.

**Table 1.**
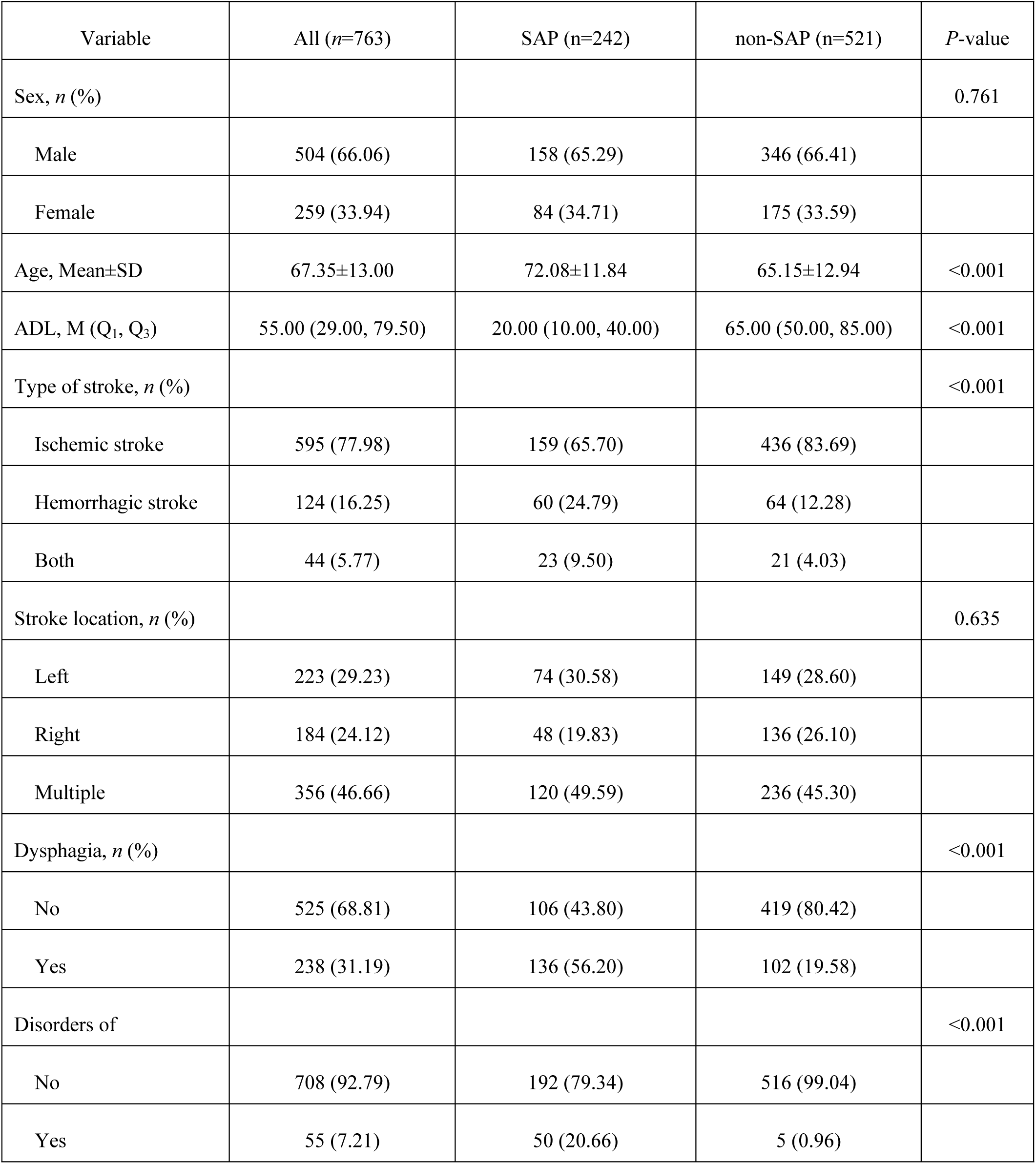
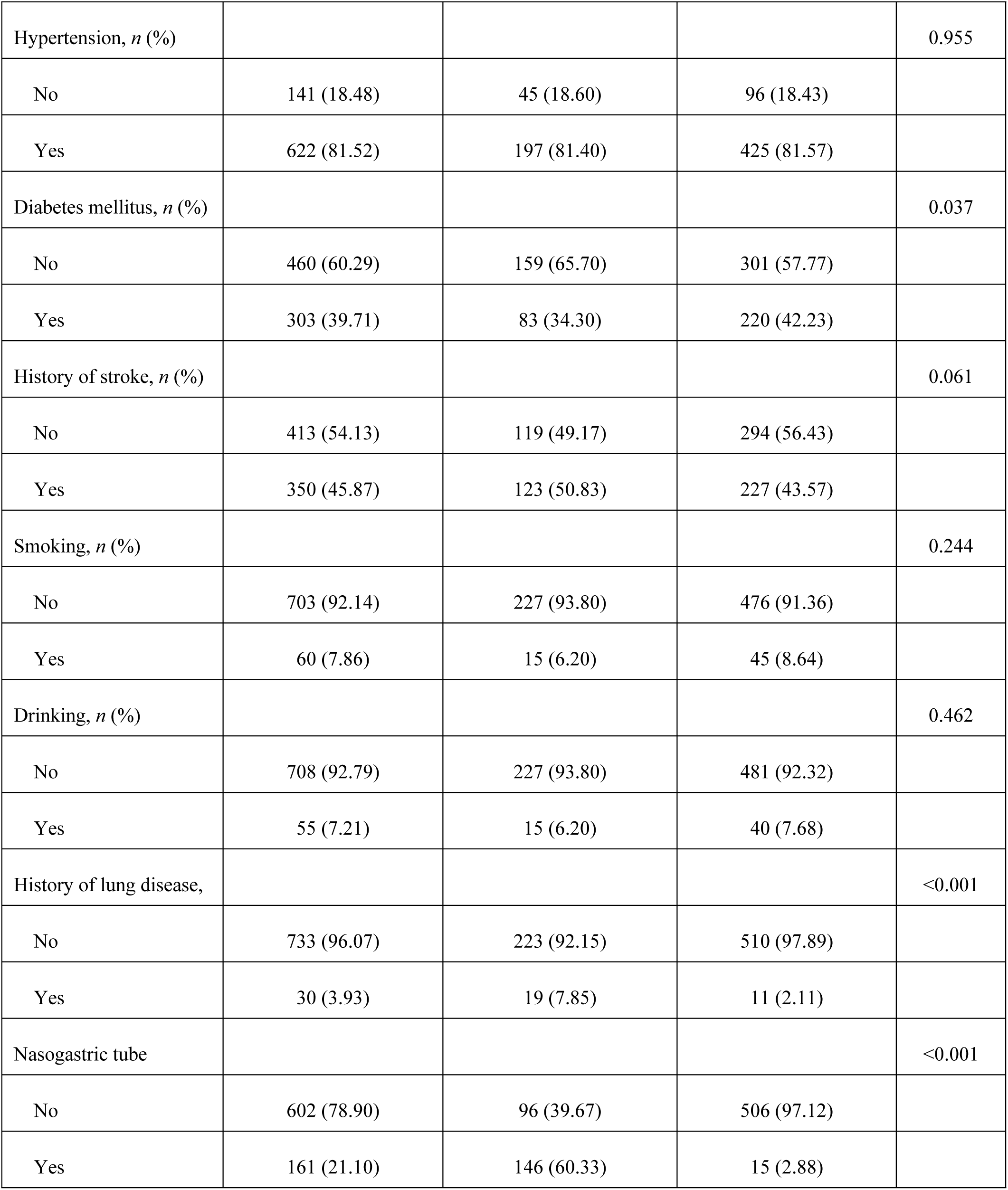
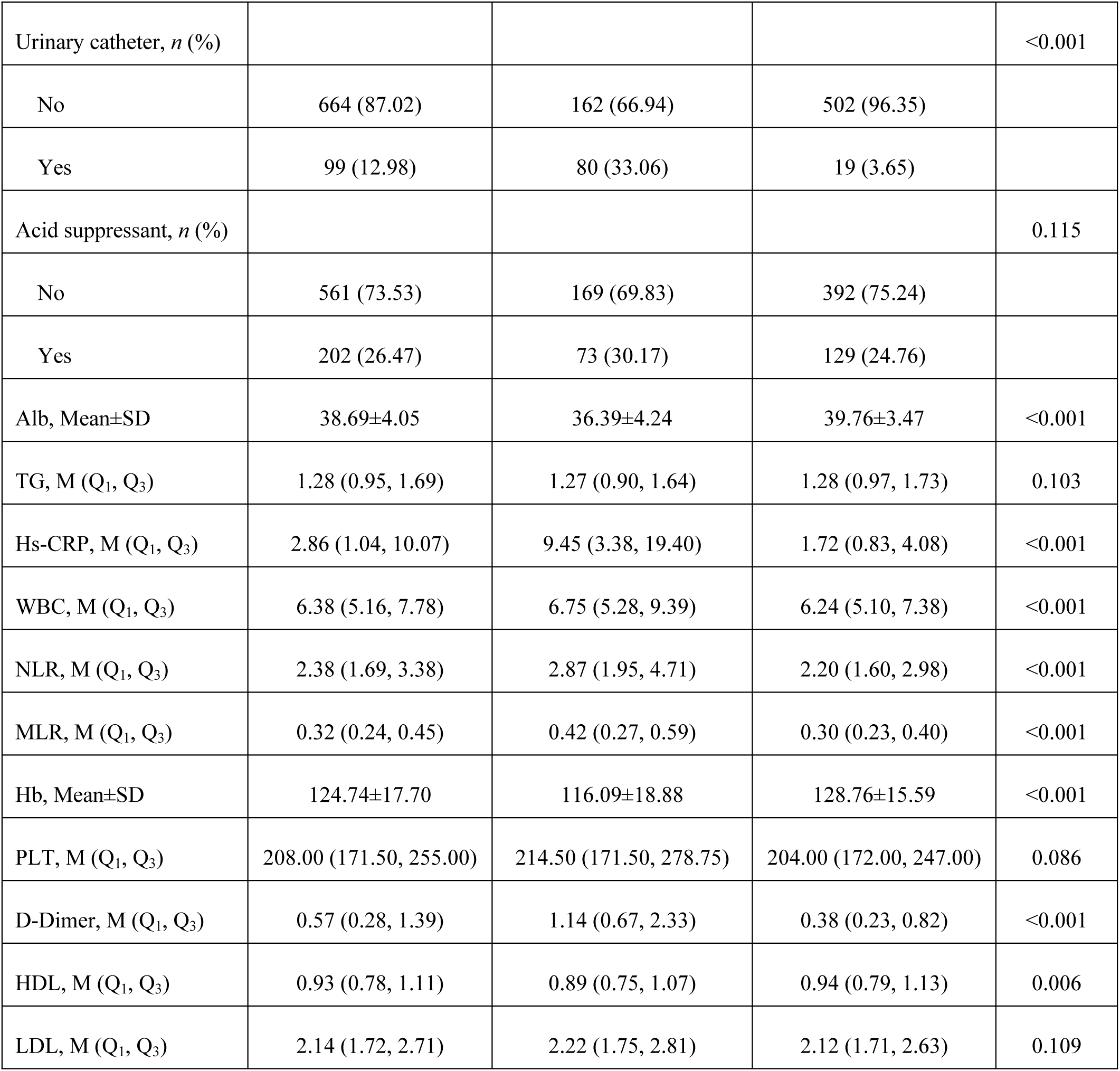
Comparison between non-SAP and SAP groups.

| Variable | All (n=763) | SAP (n=242) | non-SAP (n=521) | P-value |
| --- | --- | --- | --- | --- |
| Sex, *n* (%) |  |  |  | 0.761 |
| Male | 504 (66.06) | 158 (65.29) | 346 (66.41) |  |
| Female | 259 (33.94) | 84 (34.71) | 175 (33.59) |  |
| Age, Mean±SD | 67.35±13.00 | 72.08±11.84 | 65.15±12.94 | <0.001 |
| ADL, M (Q<sub>1</sub>, Q<sub>3</sub>) | 55.00 (29.00, 79.50) | 20.00 (10.00, 40.00) | 65.00 (50.00, 85.00) | <0.001 |
| Type of stroke, *n* (%) |  |  |  | <0.001 |
| Ischemic stroke | 595 (77.98) | 159 (65.70) | 436 (83.69) |  |
| Hemorrhagic stroke | 124 (16.25) | 60 (24.79) | 64 (12.28) |  |
| Both | 44 (5.77) | 23 (9.50) | 21 (4.03) |  |
| Stroke location, *n* (%) |  |  |  | 0.635 |
| Left | 223 (29.23) | 74 (30.58) | 149 (28.60) |  |
| Right | 184 (24.12) | 48 (19.83) | 136 (26.10) |  |
| Multiple | 356 (46.66) | 120 (49.59) | 236 (45.30) |  |
| Dysphagia, *n* (%) |  |  |  | <0.001 |
| No | 525 (68.81) | 106 (43.80) | 419 (80.42) |  |
| Yes | 238 (31.19) | 136 (56.20) | 102 (19.58) |  |
| Disorders of |  |  |  | <0.001 |
| No | 708 (92.79) | 192 (79.34) | 516 (99.04) |  |
| Yes | 55 (7.21) | 50 (20.66) | 5 (0.96) |  |
| Hypertension, *n* (%) |  |  |  | 0.955 |
| No | 141 (18.48) | 45 (18.60) | 96 (18.43) |  |
| Yes | 622 (81.52) | 197 (81.40) | 425 (81.57) |  |
| Diabetes mellitus, *n* (%) |  |  |  | 0.037 |
| No | 460 (60.29) | 159 (65.70) | 301 (57.77) |  |
| Yes | 303 (39.71) | 83 (34.30) | 220 (42.23) |  |
| History of stroke, *n* (%) |  |  |  | 0.061 |
| No | 413 (54.13) | 119 (49.17) | 294 (56.43) |  |
| Yes | 350 (45.87) | 123 (50.83) | 227 (43.57) |  |
| Smoking, *n* (%) |  |  |  | 0.244 |
| No | 703 (92.14) | 227 (93.80) | 476 (91.36) |  |
| Yes | 60 (7.86) | 15 (6.20) | 45 (8.64) |  |
| Drinking, *n* (%) |  |  |  | 0.462 |
| No | 708 (92.79) | 227 (93.80) | 481 (92.32) |  |
| Yes | 55 (7.21) | 15 (6.20) | 40 (7.68) |  |
| History of lung disease, |  |  |  | <0.001 |
| No | 733 (96.07) | 223 (92.15) | 510 (97.89) |  |
| Yes | 30 (3.93) | 19 (7.85) | 11 (2.11) |  |
| Nasogastric tube |  |  |  | <0.001 |
| No | 602 (78.90) | 96 (39.67) | 506 (97.12) |  |
| Yes | 161 (21.10) | 146 (60.33) | 15 (2.88) |  |
| Urinary catheter, *n* (%) |  |  |  | <0.001 |
| No | 664 (87.02) | 162 (66.94) | 502 (96.35) |  |
| Yes | 99 (12.98) | 80 (33.06) | 19 (3.65) |  |
| Acid suppressant, *n* (%) |  |  |  | 0.115 |
| No | 561 (73.53) | 169 (69.83) | 392 (75.24) |  |
| Yes | 202 (26.47) | 73 (30.17) | 129 (24.76) |  |
| Alb, Mean±SD | 38.69±4.05 | 36.39±4.24 | 39.76±3.47 | <0.001 |
| TG, M (Q<sub>1</sub>, Q<sub>3</sub>) | 1.28 (0.95, 1.69) | 1.27 (0.90, 1.64) | 1.28 (0.97, 1.73) | 0.103 |
| Hs-CRP, M (Q<sub>1</sub>, Q<sub>3</sub>) | 2.86 (1.04, 10.07) | 9.45 (3.38, 19.40) | 1.72 (0.83, 4.08) | <0.001 |
| WBC, M (Q<sub>1</sub>, Q<sub>3</sub>) | 6.38 (5.16, 7.78) | 6.75 (5.28, 9.39) | 6.24 (5.10, 7.38) | <0.001 |
| NLR, M (Q<sub>1</sub>, Q<sub>3</sub>) | 2.38 (1.69, 3.38) | 2.87 (1.95, 4.71) | 2.20 (1.60, 2.98) | <0.001 |
| MLR, M (Q<sub>1</sub>, Q<sub>3</sub>) | 0.32 (0.24, 0.45) | 0.42 (0.27, 0.59) | 0.30 (0.23, 0.40) | <0.001 |
| Hb, Mean±SD | 124.74±17.70 | 116.09±18.88 | 128.76±15.59 | <0.001 |
| PLT, M (Q<sub>1</sub>, Q<sub>3</sub>) | 208.00 (171.50, 255.00) | 214.50 (171.50, 278.75) | 204.00 (172.00, 247.00) | 0.086 |
| D-Dimer, M (Q<sub>1</sub>, Q<sub>3</sub>) | 0.57 (0.28, 1.39) | 1.14 (0.67, 2.33) | 0.38 (0.23, 0.82) | <0.001 |
| HDL, M (Q<sub>1</sub>, Q<sub>3</sub>) | 0.93 (0.78, 1.11) | 0.89 (0.75, 1.07) | 0.94 (0.79, 1.13) | 0.006 |
| LDL, M (Q<sub>1</sub>, Q<sub>3</sub>) | 2.14 (1.72, 2.71) | 2.22 (1.75, 2.81) | 2.12 (1.71, 2.63) | 0.109 |

### 3.3 Correlation Analysis of Variables and Variable Selection

To explore the relationship between variables, Spearman’s rank correlation coefficient was used to assess the linear relationship between variables (Fig 2).We performed the Lasso regression method to screen the 27 included variables. The optimal λ value corresponding to one standard error of the minimum mean square error was determined through 10-fold cross-validation. The results indicated that when log(λ) reached one standard error of the minimum mean square error, the number of model variables was reduced to 6, with the optimal λ value determined to be 0.039 through cross-validation. The final variables included Nasogastric tube therapy, Age, ADL, Alb, Hs-CRP, Hb. The procedure for selecting variables by Lasso regression is shown in Fig 3A and Fig 3B.

**Fig 2.**
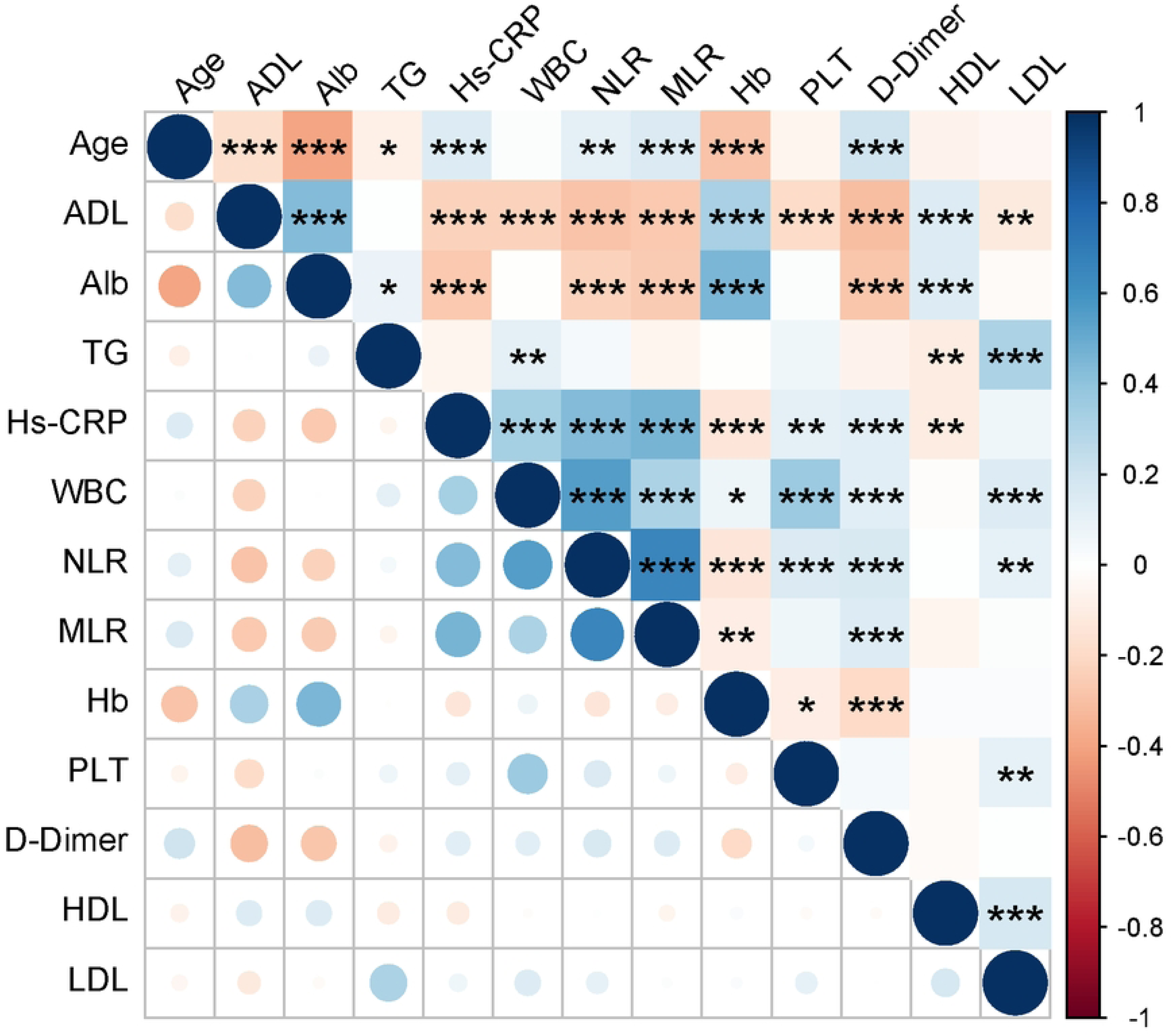
Correlation between variables.

**Fig 3.**
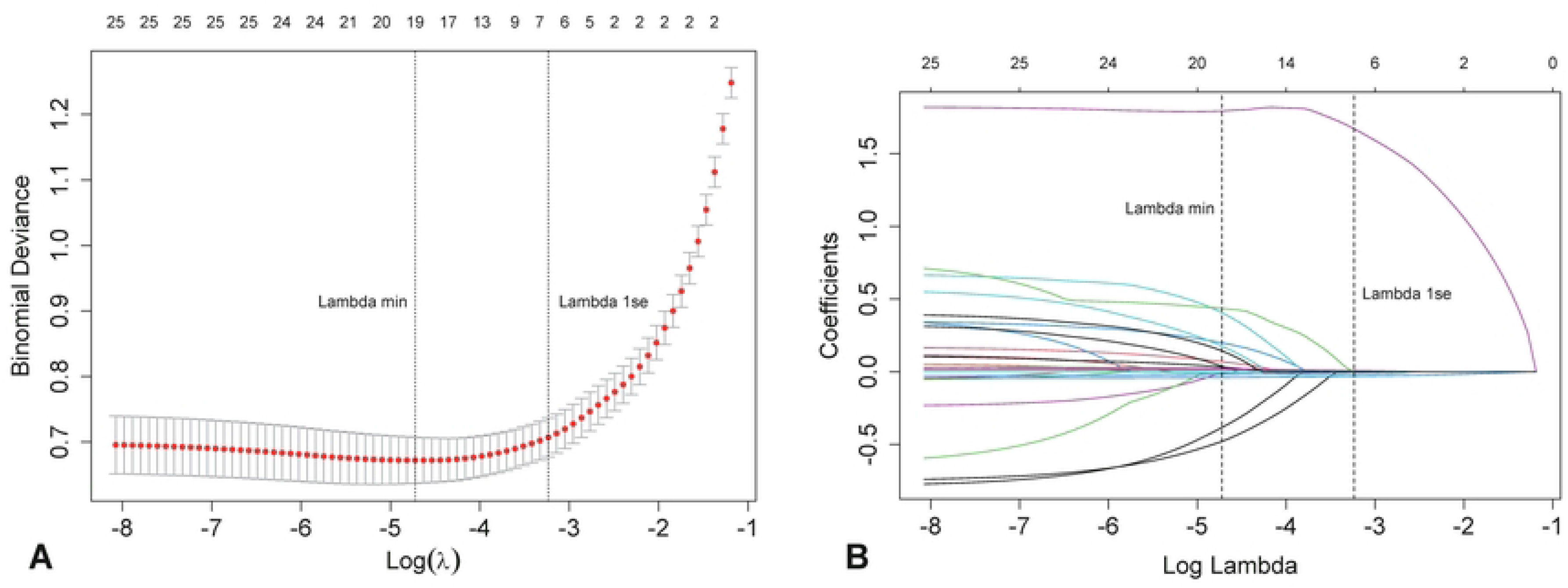
Lasso regression procedure for screening variables. (A) Deviation at different values of lambda. (B) Plot of lambda versus partial regression coefficients.

### 3.4 Model construction and evaluation

Five machine learning models were constructed using the six identified variables. In the training set, 5-fold cross-validation was employed as a method for data resampling to ensure the model’s performance on unseen data. Additionally, grid search was utilized to optimize the key hyperparameters, which included the number of features, the number of trees, the minimum number of samples, the maximum depth of trees, and the learning rate. The validation set was utilized to assess the model’s performance. The ROC curves for the five machine learning models in the validation set revealed that the XGBoost model achieved the highest AUC value of 0.926 (Fig. 4). The calibration curves for each model indicated that the average predicted probability aligned with the actual occurrence probability for all models except for the decision tree (DT) and support vector machine (SVM) models (Fig. 5). Decision curve analysis (DCA) demonstrated that all models maintained good net benefits within a certain threshold probability, indicating high clinical utility value (Fig. 6). The XGBoost model also demonstrated superior predictive performance and practical utility in both the calibration curve and DCA. A comparison of the performance indicators for the five machine learning models in the validation set showed that the XGBoost model continued to excel, with an AUC of 0.926, an accuracy of 0.914, and an F1 score of 0.889. Additional model indicators are presented in Table 2.

**Fig 4.**
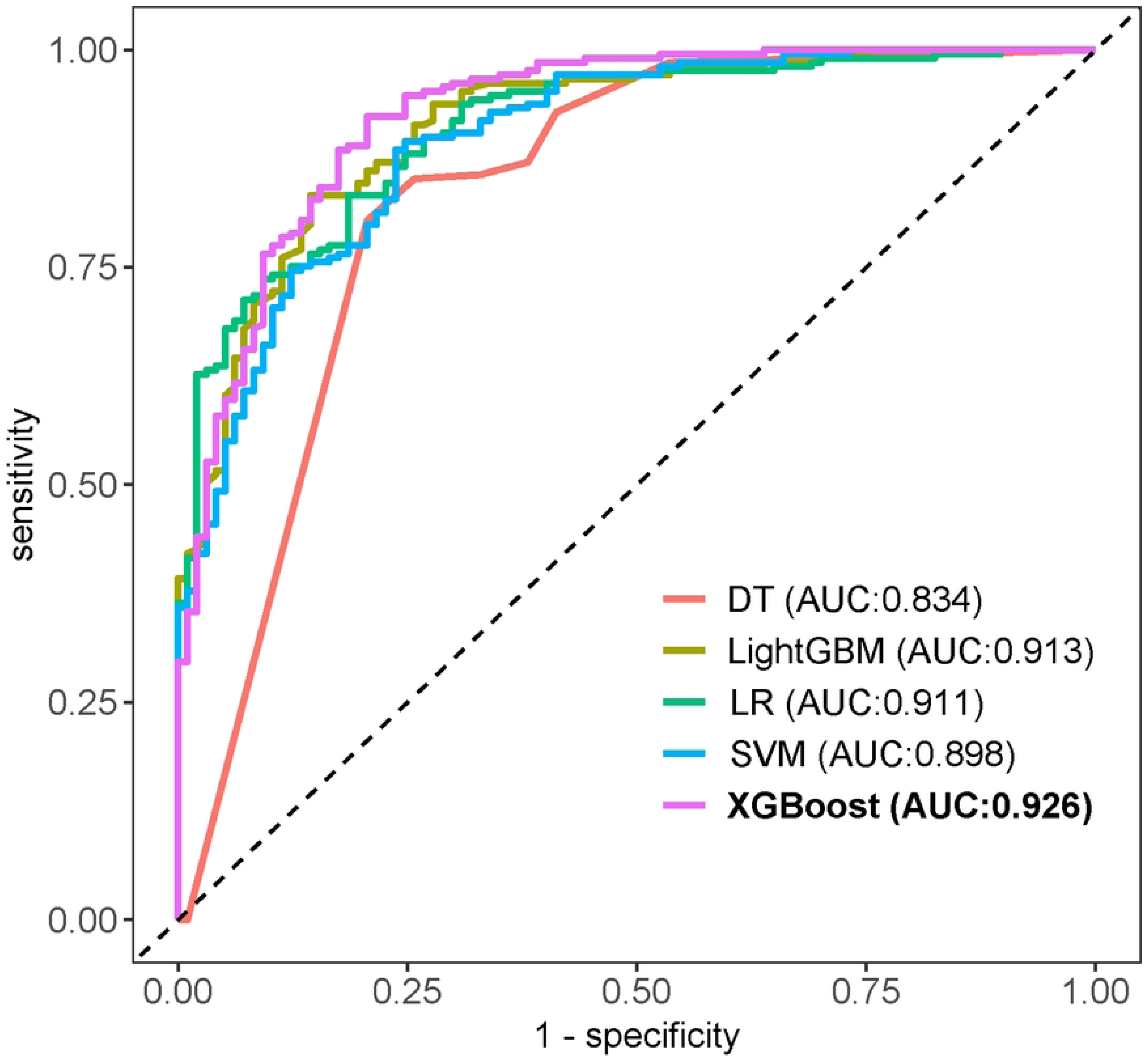
ROC curves of the five machine learning models.

**Fig 5.**
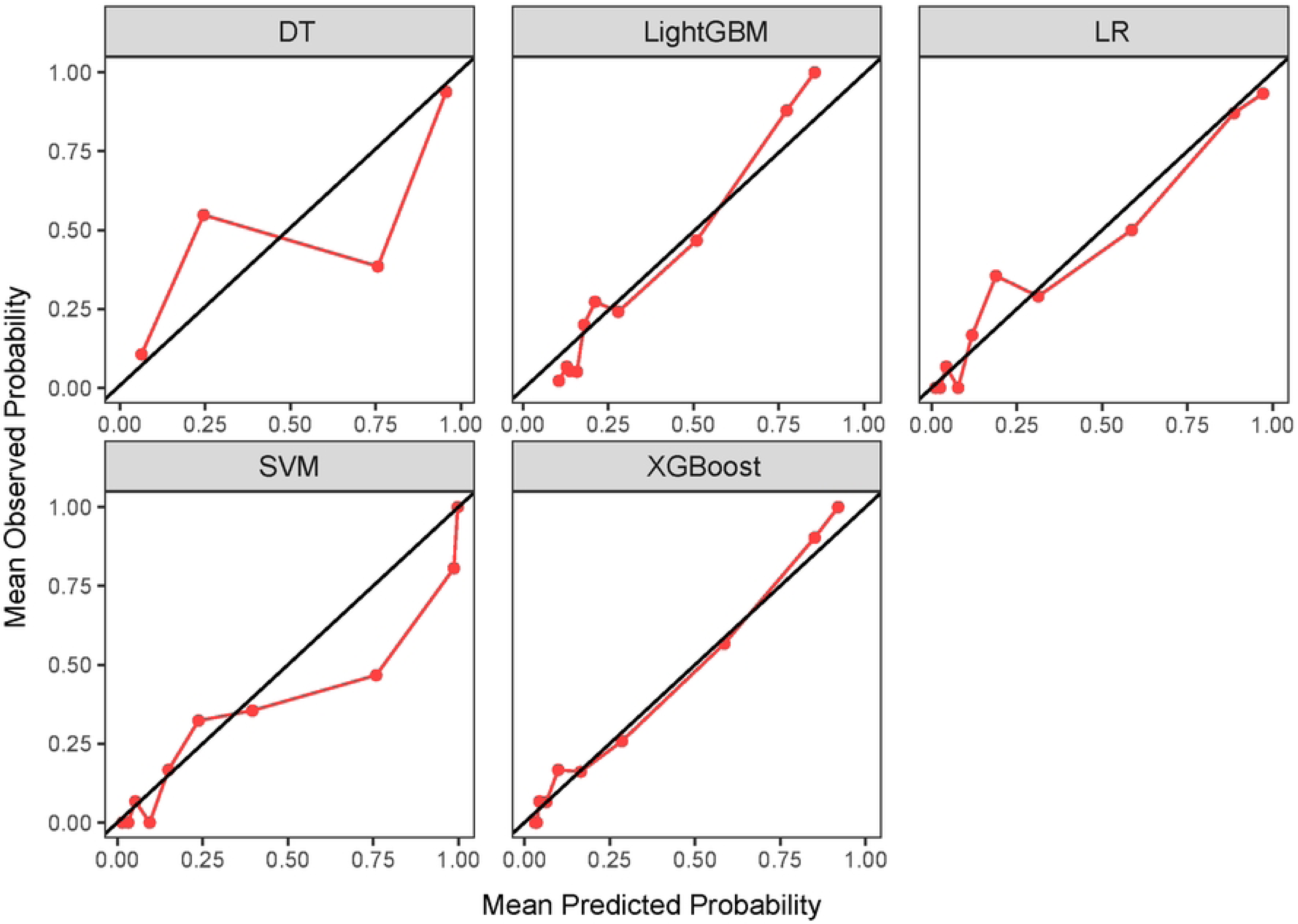
Calibration curves of five machine learning models.

**Fig 6.**
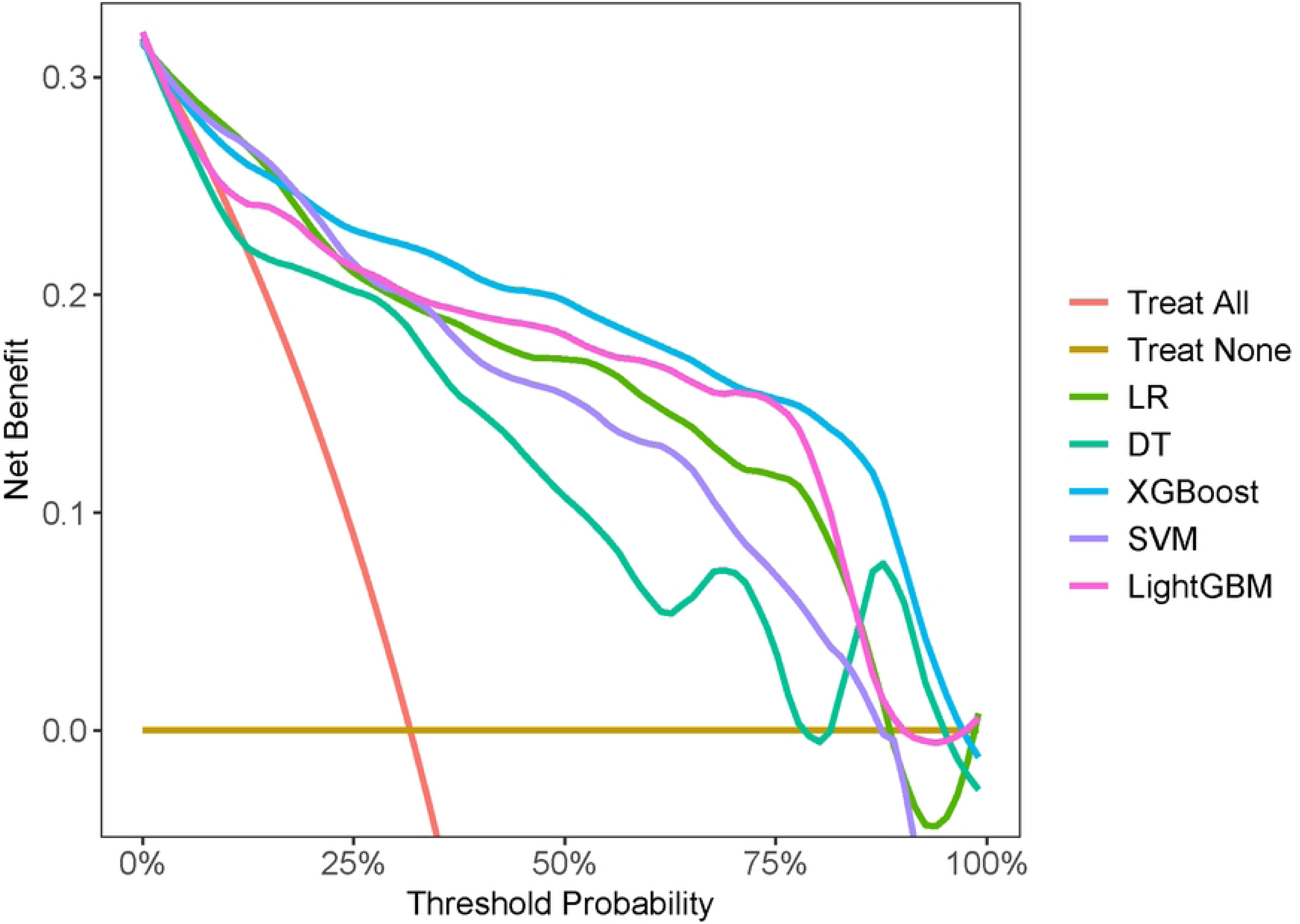
DCA for five machine learning models.

**Table 2.** Comparison of predictive performance metrics for various models in the validation set.

|  | DT | LR | XGBoost | SVM | LightGBM |
| --- | --- | --- | --- | --- | --- |
| Accuracy | 0.817 | 0.833 | 0.853 | 0.781 | 0.840 |
| Kappa | 0.584 | 0.614 | 0.670 | 0.529 | 0.635 |
| Sensitivity | 0.852 | 0.880 | 0.866 | 0.775 | 0.871 |
| Specificity | 0.742 | 0.732 | 0.825 | 0.794 | 0.773 |
| PPV | 0.877 | 0.876 | 0.914 | 0.890 | 0.892 |
| NPV | 0.699 | 0.740 | 0.741 | 0.621 | 0.735 |
| MCC | 0.585 | 0.614 | 0.673 | 0.539 | 0.636 |
| J-index | 0.594 | 0.612 | 0.691 | 0.569 | 0.644 |
| Balanced Accuracy | 0.797 | 0.806 | 0.845 | 0.784 | 0.822 |
| Detection Prevalence | 0.663 | 0.686 | 0.647 | 0.595 | 0.667 |
| Precision | 0.877 | 0.876 | 0.914 | 0.890 | 0.892 |
| Recall | 0.852 | 0.880 | 0.866 | 0.775 | 0.871 |
| F-measure | 0.864 | 0.878 | 0.889 | 0.829 | 0.881 |
| AUC | 0.834 | 0.911 | 0.926 | 0.898 | 0.913 |

PPV, positive predictive value; NPV, negative predictive value; MCC, matthews correlation coefficient

### 3.5 Interpreting the XGBoost model using SHAP

The importance ranking plot of the variables showed that ADL had the highest mean SHAP value and the strongest predictive performance (Fig 7). To analyze the positive and negative correlation between the variable and the target outcome, we drew a beeswarm in which the color depth reflected the value of the variable. Taking the first behavior as an example, the low ADL level had a negative effect on the outcome prediction. High ADL level was a positive predictor of outcome (Fig 8). Finally, the SHAP value is used to show the influence of different variables in a sample on the prediction result of SAP occurrence risk. The value in the Fig represents the contribution degree of each feature to the output result of the model. A positive number indicates that when the feature increases, the predictive value of the model will also increase. The opposite is true for negative numbers (Fig 9).

**Fig 7.**
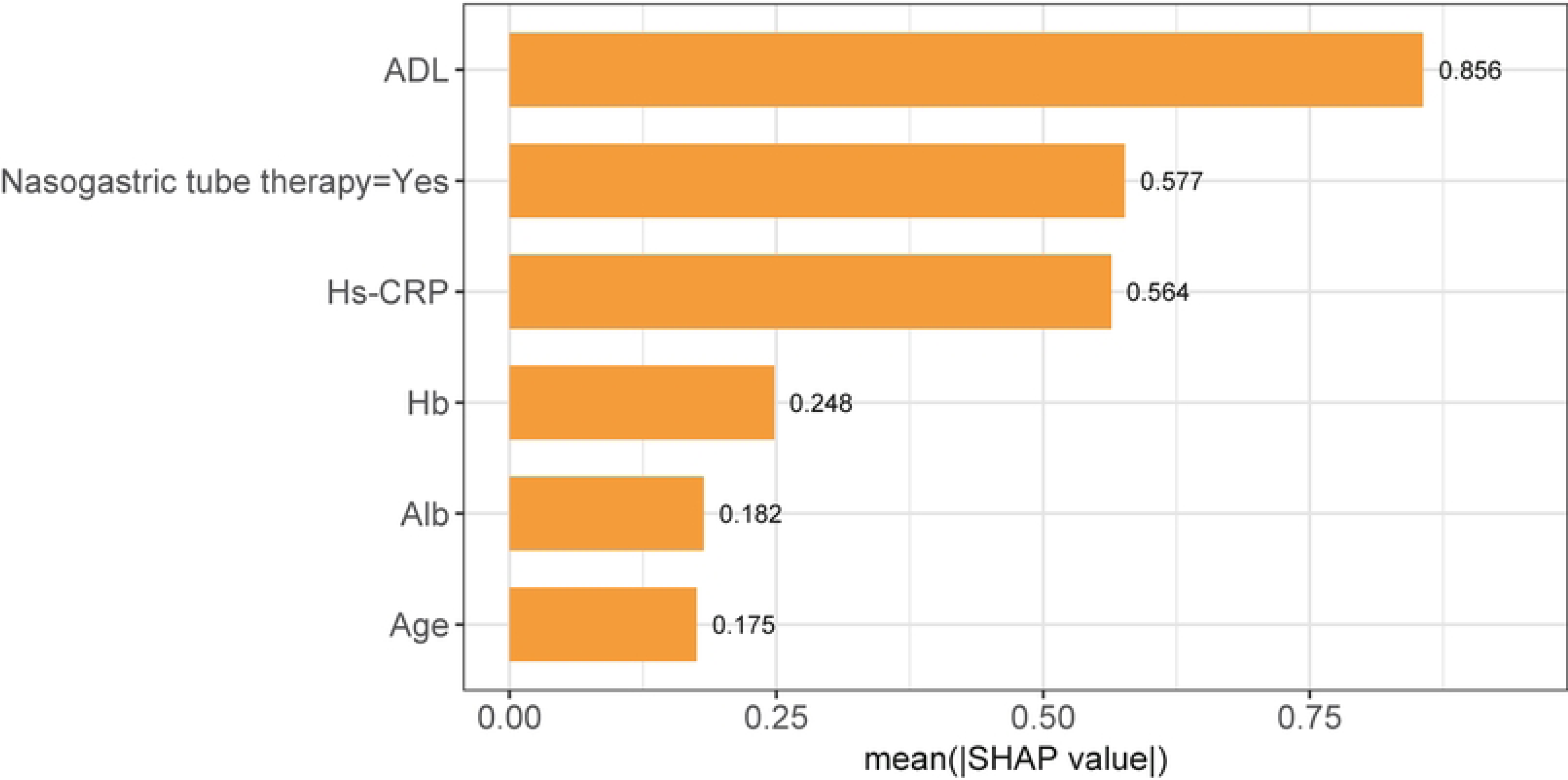
The importance ranking of variables was analyzed based on SHAP algorithm.

**Fig 8.**
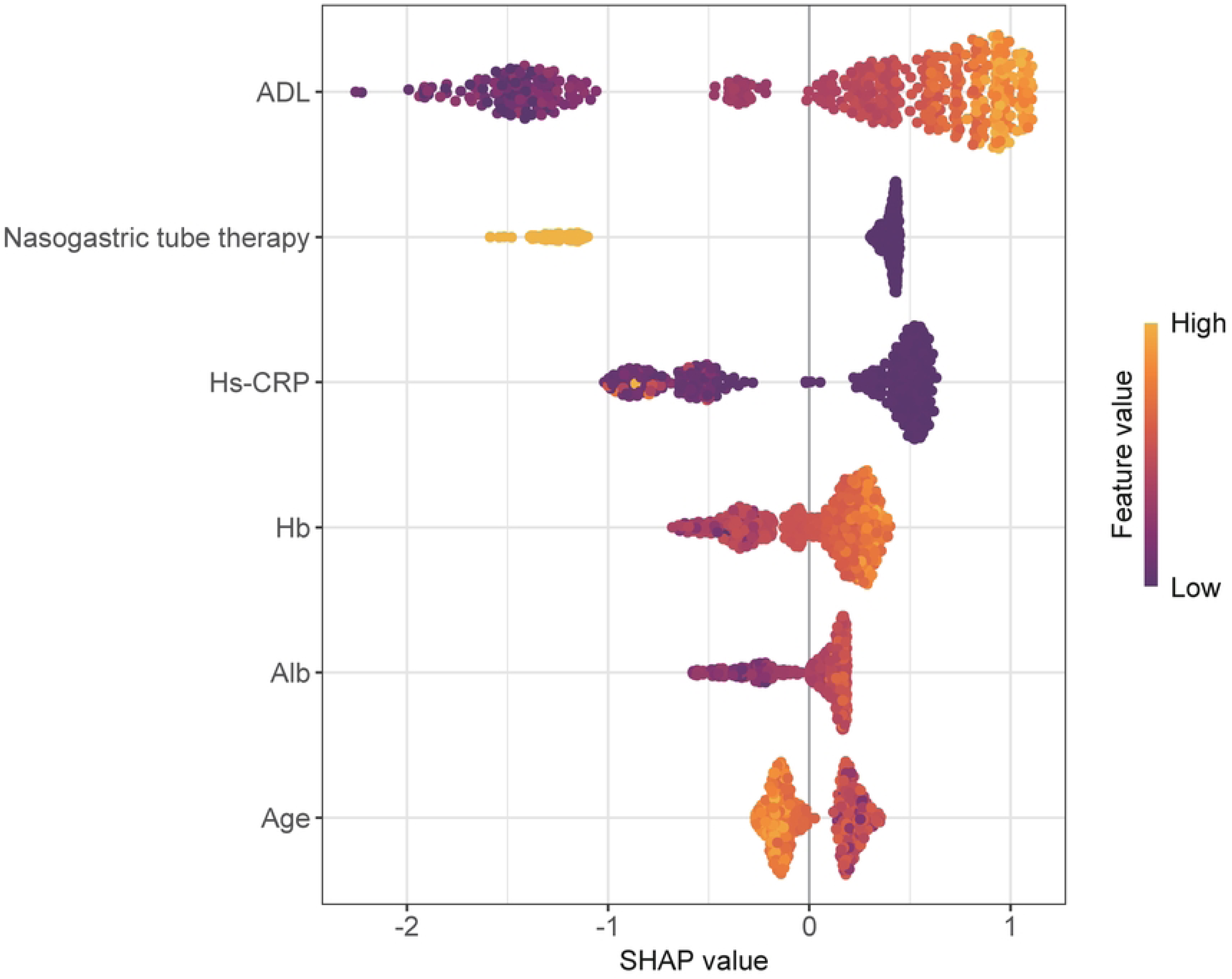
The beeswarm of the XGBoost model was analyzed based on SHAP algorithm.

**Fig 9.**
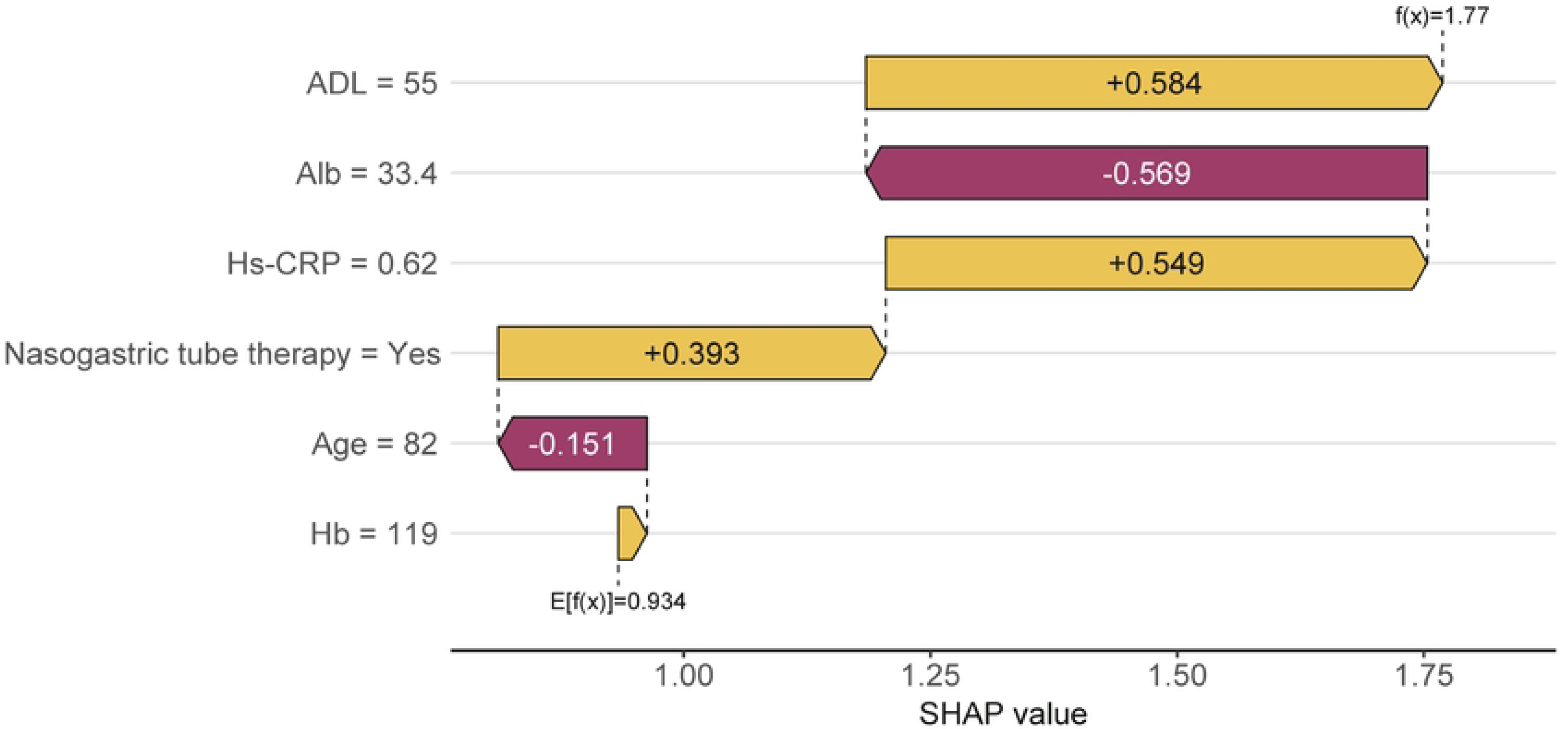
The risk of SAP in a single sample was analyzed based on SHAP algorithm.

## 4. Discussion

In this study, five machine learning methods were used to construct a risk prediction model for SAP in stroke patients based on six predictors, including Nasogastric tube therapy, Age, ADL, Alb, Hs-CRP, Hb. Among them, the XGBoost model validation set showed good discrimination and calibration. The prediction performance of the models was better than those of previous studies [10–12]. Additionally, the model was visualized using a nomogram, making the risk scoring more intuitive and quantifiable.

SHAP algorithm results showed that the lower the ADL score, the higher the risk of SAP. It was found that patients with SAP tended to have longer hospital stays and lower ADL scores compared with non-SAP patients[18]. A low ADL score typically indicates a limited ability to perform self-care, potentially leading to prolonged bed rest, which indirectly increases the risk of pulmonary infections[19]. Furthermore, low ADL scores may correlate with poor nutritional status, compromising the immune system and making patients more prone to infections. Early rehabilitation exercises can enhance the self-care abilities of stroke patients, reducing the risk of lung infections, and clinical practitioners should promptly intervene in patients with limited mobility.

In this study, indwelling nasogastric tube was a risk factor for SAP in stroke patients. This result is consistent with reports in the existing study[20].Brogan found that nasogastric tubes were a stronger predictor of SAP than dysphagia[21]. Research indicates that during nasogastric tube feeding, the gastric volume significantly expands, potentially causing gastric muscle spasms and food accumulation. In the mean time, patients with nasogastric tubes face increased risks of elevated intracranial pressure, vomiting, and food reflux, collectively heightening the probability of SAP[22]. However, a study on acute stroke patients found that placing a nasogastric tube within 48 hours of onset did not increase the incidence of SAP, mortality, or adverse functional outcomes[23], contradicting our findings. Thus, the relationship between the duration of nasogastric tube indwelling and SAP remains to be studied.

This finding suggests that older stroke patients are more susceptible to SAP, consistent with previous studies[11,12]. As individuals age, their physiological functions and immune system capabilities deteriorate, weakening respiratory defenses. Additionally, older stroke patients often exhibit diminished swallowing and coughing reflexes, making them more vulnerable to pulmonary infections post-stroke[24].

Among the laboratory examination indicators, high Hs-CRP level was a factor affecting the occurrence of SAP, which was consistent with the results of previous studies[25]. Hs-CRP represents an acute phase reactive protein produced by the liver, falling under the category of C-reactive protein (CRP). Hs-CRP demonstrates superior sensitivity and early discriminative capabilities, making it detectable with high efficiency during the initial phases of inflammation or at minimal concentrations[26]. Research indicates that alterations in serum Hs-CRP levels among patients following cerebral infarction are strongly correlated with the occurrence of secondary pulmonary infections. As Hs-CRP levels rise, the inflammatory response becomes more pronounced[27]. Another effective predictor of SAP is Hb, and the decrease in Hb level usually means the patient is at risk of anemia, which can significantly increase the mortality and the risk of pneumonia[28]. Anemia is associated with compromised immune system function, thereby compromising the patient’s immune response and rendering stroke patients more vulnerable to bacterial or viral pathogens[29]. Several prior studies have established that stroke patients with reduced serum Alb levels are at an elevated risk of contracting infections or pneumonia while hospitalized[30–32], one possible reason is that Alb has anti-inflammatory, anti-oxidative, anticoagulant effects as well as regulation of microvascular permeability; therefore, low Alb level is a marker of systemic inflammatory response[32]. Furthermore, reduced levels of Alb can precipitate malnutrition in patients, which in turn diminishes the patient’s overall health and impairs their resistance to infection, consequently elevating the risk of pneumonia[33].

Machine learning algorithms demonstrate substantial superiority over traditional regression techniques in handling high-dimensional data, intricate relationships, and feature selection, while also enhancing prediction accuracy. In this study, XGBoost exhibited robust predictive capabilities, currently standing as one of the most widely used machine learning algorithms. It offers high accuracy, incorporates regularization, and boasts both high prediction performance and interpretability, hence holding great promise for application in risk prediction and various other domains[34–36]. In order to achieve the best accuracy on large modern datasets, it is often necessary to rely on complex models. However, complex models are often difficult to balance the contradiction between model accuracy and interpretability. Based on this, the SHAP framework provides a powerful tool for model interpretation that can help us balance model accuracy and interpretability and make better use of complex models in practical applications[37].

## 5. Limitations

Despite the robust performance of our model in various aspects, there are limitations. First, this study is a single-center, retrospective study, with data collection primarily relying on nursing and physician records, which may result in limited data and potential information bias. Secondly, our model has not yet undergone external validation in other populations, so its generalizability remains to be further examined. Additionally, the model depends on accurate data input, and data integrity in practical applications could affect the predictive performance. Future research should aim to optimize the model further, incorporating more potential predictive factors and exploring advanced machine learning algorithms.

## 6. Conclusion

The models developed in this study demonstrate strong predictive capabilities and clinical utility in assessing the risk of stroke-associated pneumonia in stroke patients, with the XGBoost model achieving the highest performance. Furthermore, the SHAP framework has been instrumental in elucidating the models, identifying the critical factors influencing the onset of stroke-associated pneumonia, and has furnished crucial insights for clinical decision-making. Our findings provide a new tool for SAP prediction, which may support clinical decision-making and improve patient outcomes.

## Data Availability

All relevant data are within the manuscript and its Supporting Information files.

## Author Contributions

Chunbiao Li: Writing – original draft, Writing – review & editing, Conceptualization, Software;

Formal Analysis, Visualization

Ting Wang: Writing – review & editing, Conceptualization, Methodology, Funding acquisition;

Juan Yuan: Writing – review & editing, Conceptualization, Methodology; Supervision;

Linli Yuan: Writing – review & editing, Funding acquisition, Data curation

Min You: Writing – review & editing, Supervision, Funding acquisition, Data curation.

## Acknowledgments

The authors are very grateful to the authority of the Second Affiliated Hospital of Anhui University of Traditional Chinese Medicine for allowing us to use these data for the construction of the model. The authors would like to thank Dajin Li and Yi Liu for their valuable help.

## Supporting information

S1 Fig. Graphical representation of cross validation of the optimal hyperparameters of the XGBoost model

S2 Fig. Confusion matrix of the XGBoost model in the training and validation sets

S3 Fig. Importance ranking of variables in the XGBoost model

S4 Fig. Performance of five machine learning models in the validation set

S5 Fig. Comparison of five machine learning models with five-fold cross-validation (AUC as evaluation index)

S6 Fig. Dependence plot of ADL

S7 Fig. Dependence plot of Hs-CRP

S8 Fig. Dependence plot of Age

S9 Fig. Dependence plot of Nasogastric tube therapy

S10 Fig. Dependence plot of Alb

S11 Fig. Dependence plot of Hb

## References

1. Feigin VL, Stark BA, Johnson CO, Roth GA, Bisignano C, Abady GG, et al. Global, regional, and national burden of stroke and its risk factors, 1990–2019: a systematic analysis for the Global Burden of Disease Study 2019. Lancet Neurol. 2021;20: 795–820. doi:10.1016/S1474-4422(21)00252-0

2. Smith CJ, Kishore AK, Vail A, Chamorro A, Garau J, Hopkins SJ, et al. Diagnosis of Stroke-Associated Pneumonia: Recommendations from the Pneumonia in Stroke Consensus Group. Stroke. 2015;46: 2335–2340. doi:10.1161/STROKEAHA.115.009617

3. Ji R, Wang D, Shen H, Pan Y, Liu G, Wang P, et al. Interrelationship among common medical complications after acute stroke: pneumonia plays an important role. Stroke. 2013;44: 3436–3444. doi:10.1161/STROKEAHA.113.001931

4. Smith CJ, Bray BD, Hoffman A, Meisel A, Heuschmann PU, Wolfe CDA, et al. Can a Novel Clinical Risk Score Improve Pneumonia Prediction in Acute Stroke Care? A UK Multicenter Cohort Study. J Am Heart Assoc. 2015;4: e001307. doi:10.1161/JAHA.114.001307

5. Teh WH, Smith CJ, Barlas RS, Wood AD, Bettencourt-Silva JH, Clark AB, et al. Impact of stroke-associated pneumonia on mortality, length of hospitalization, and functional outcome. Acta Neurol Scand. 2018;138: 293–300. doi:10.1111/ane.12956

6. Yu Y-J, Weng W-C, Su F-C, Peng T-I, Chien Y-Y, Wu C-L, et al. Association between pneumonia in acute stroke stage and 3-year mortality in patients with acute first-ever ischemic stroke. J Clin Neurosci. 2016;33: 124–128. doi:10.1016/j.jocn.2016.02.039

7. Houck PM, Bratzler DW, Nsa W, Ma A, Bartlett JG. Timing of antibiotic administration and outcomes for Medicare patients hospitalized with community-acquired pneumonia. Arch Intern Med. 2004;164: 637–644. doi:10.1001/archinte.164.6.637

8. Vermeij J-D, Westendorp WF, Dippel DW, van de Beek D, Nederkoorn PJ. Antibiotic therapy for preventing infections in people with acute stroke. Cochrane Database Syst Rev. 2018;1: CD008530. doi:10.1002/14651858.CD008530.pub3

9. Ranstam J, Cook JA, Collins GS. Clinical prediction models. Br J Surg. 2016;103: 1886. doi:10.1002/bjs.10242

10. Kwon H-M, Jeong S-W, Lee S-H, Yoon B-W. The pneumonia score: A simple grading scale for prediction of pneumonia after acute stroke. Am J Infect Control. 2006;34: 64–68. doi:10.1016/j.ajic.2005.06.011

11. Hoffmann S, Malzahn U, Harms H, Koennecke H-C, Berger K, Kalic M, et al. Development of a Clinical Score (A2DS2) to Predict Pneumonia in Acute Ischemic Stroke. Stroke. 2012;43: 2617–2623. doi:10.1161/STROKEAHA.112.653055

12. Ji R, Shen H, Pan Y, Wang P, Liu G, Wang Y, et al. Novel Risk Score to Predict Pneumonia After Acute Ischemic Stroke. Stroke. 2013;44: 1303–1309. doi:10.1161/STROKEAHA.111.000598

13. Gu H, Zhou Z, Zhang Z, Zhou Q. Clinical Prediction Models: Basic Concepts, Application Scenarios, and Research Strategies. Chin J Evid Based Cardiovasc Med. 2018;10: 1454–1456+1462.

14. Collin C, Wade DT, Davies S, Horne V. The Barthel ADL Index: A reliability study. Int Disabil Stud. 1988;10: 61–63. doi:10.3109/09638288809164103

15. Horan TC, Andrus M, Dudeck MA. CDC/NHSN surveillance definition of health care-associated infection and criteria for specific types of infections in the acute care setting. Am J Infect Control. 2008;36: 309–332. doi:10.1016/j.ajic.2008.03.002

16. Riley RD, Ensor J, Snell KIE, Harrell FE, Martin GP, Reitsma JB, et al. Calculating the sample size required for developing a clinical prediction model. BMJ. 2020; m441. doi:10.1136/bmj.m441

17. CAO Y, ZENG X, ZHAO X, PENG M. Risk prediction models for stroke-associated pneumonia: a systematic review. Chin J Evid-Based Med. 2023;23: 1259–1268.

18. Maeshima S, Osawa A, Hayashi T, Tanahashi N. Elderly Age, Bilateral Lesions, and Severe Neurological Deficit Are Correlated with Stroke-associated Pneumonia. J Stroke Cerebrovasc Dis. 2014;23: 484–489. doi:10.1016/j.jstrokecerebrovasdis.2013.04.004

19. Matsusaka K, Kawakami G, Kamekawa H, Momma H, Nagatomi R, Itoh J, et al. Pneumonia risks in bedridden patients receiving oral care and their screening tool: Malnutrition and urinary tract infection-induced inflammation. Geriatr Gerontol Int. 2018;18: 714–722. doi:10.1111/ggi.13236

20. Schwarz M, Coccetti A, Murdoch A, Cardell E. The impact of aspiration pneumonia and nasogastric feeding on clinical outcomes in stroke patients: A retrospective cohort study. J Clin Nurs. 2018;27. doi:10.1111/jocn.13922

21. Brogan E, Langdon C, Brookes K, Budgeon C, Blacker D. Respiratory infections in acute stroke: nasogastric tubes and immobility are stronger predictors than dysphagia. Dysphagia. 2014;29: 340–345. doi:10.1007/s00455-013-9514-5

22. Wang Q, Liu Y, Han L, He F, Cai N, Zhang Q, et al. Risk factors for acute stroke-associated pneumonia and prediction of neutrophil-to-lymphocyte ratios. Am J Emerg Med. 2021;41: 55–59. doi:10.1016/j.ajem.2020.12.036

23. Kalra L, Hodsoll J, Irshad S, Smithard D, Manawadu D, STROKE-INF Investigators. Association between nasogastric tubes, pneumonia, and clinical outcomes in acute stroke patients. Neurology. 2016;87: 1352–1359. doi:10.1212/WNL.0000000000003151

24. Patel UK, Kodumuri N, Dave M, Lekshminarayanan A, Khan N, Kavi T, et al. Stroke-Associated Pneumonia: A Retrospective Study of Risk Factors and Outcomes. The Neurologist. 2020;25: 39–48. doi:10.1097/NRL.0000000000000269

25. Huang L, Zhang R, Ji J, Long F, Wang Y, Lu J, et al. Hypersensitive C-reactive protein-albumin ratio is associated with stroke-associated pneumonia and early clinical outcomes in patients with acute ischemic stroke. Brain Behav. 2022;12: e2675. doi:10.1002/brb3.2675

26. Banait T, Wanjari A, Danade V, Banait S, Jain J. Role of High-Sensitivity C-reactive Protein (Hs-CRP) in Non-communicable Diseases: A Review. Cureus. 2022;14: e30225. doi:10.7759/cureus.30225

27. Wang L, Wu L, Lang Y, Wu D, Chen J, Zhao W, et al. Association between high-sensitivity C-reactive protein levels and clinical outcomes in acute ischemic stroke patients treated with endovascular therapy. Ann Transl Med. 2020;8: 1379. doi:10.21037/atm-20-3820

28. Wei C -C., Zhang S -T., Tan G, Zhang S -H., Liu M. Impact of anemia on in-hospital complications after ischemic stroke. Eur J Neurol. 2018;25: 768–774. doi:10.1111/ene.13595

29. Song X, He Y, Bai J, Zhang J. A nomogram based on nutritional status and A2DS2 score for predicting stroke-associated pneumonia in acute ischemic stroke patients with type 2 diabetes mellitus: A retrospective study. Front Nutr. 2022;9: 1009041. doi:10.3389/fnut.2022.1009041

30. Yang X, Wang L, Zheng L, Wu J, Liu J, Hao Z, et al. Serum Albumin as a Potential Predictor of Pneumonia after an Acute Ischemic Stroke. Curr Neurovasc Res. 2020;17: 385–393. doi:10.2174/1567202617666200514120641

31. Huang L, Zhang R, Ji J, Long F, Wang Y, Lu J, et al. Hypersensitive C-reactive protein-albumin ratio is associated with stroke-associated pneumonia and early clinical outcomes in patients with acute ischemic stroke. Brain Behav. 2022;12: e2675. doi:10.1002/brb3.2675

32. Lin G, Hu M, Song J, Xu X, Liu H, Qiu L, et al. High Fibrinogen to Albumin Ratio: A Novel Marker for Risk of Stroke-Associated Pneumonia? Front Neurol. 2022;12: 747118. doi:10.3389/fneur.2021.747118

33. Chen L, Xu M, Huang Q, Liu Y, Ren W. Clinical significance of albumin to globulin ratio among patients with stroke-associated pneumonia. Front Nutr. 2022;9: 970573. doi:10.3389/fnut.2022.970573

34. Zhang Y, Xiang T, Wang Y, Shu T, Yin C, Li H, et al. Explainable machine learning for predicting 30-day readmission in acute heart failure patients. iScience. 2024;27: 110281. doi:10.1016/j.isci.2024.110281

35. Islam MM, Rahman MJ, Rabby MS, Alam MJ, Pollob SMAI, Ahmed NAMF, et al. Predicting the risk of diabetic retinopathy using explainable machine learning algorithms. Diabetes Metab Syndr. 2023;17: 102919. doi:10.1016/j.dsx.2023.102919

36. Hou N, Li M, He L, Xie B, Wang L, Zhang R, et al. Predicting 30-days mortality for MIMIC-III patients with sepsis-3: a machine learning approach using XGboost. J Transl Med. 2020;18: 1–14. doi:10.1186/s12967-020-02620-5

37. Lundberg SM, Lee S-I. A unified approach to interpreting model predictions. Proceedings of the 31st International Conference on Neural Information Processing Systems. Red Hook, NY, USA: Curran Associates Inc.; 2017. pp. 4768–4777.

